# State Medicaid Expansion Policies were Associated with Reduced Rates of Congenital Syphilis, 2008-2022

**DOI:** 10.1101/2025.03.02.25323181

**Authors:** Austin M Williams

## Abstract

**Objective:** To assess the relationship between adoption of Medicaid expansion policies and rates of congenital syphilis (CS) in the United States.

**Study Setting and Design:** Cases of CS have increased nearly 10-fold in the past decade. State (and D.C.) adoptions of Medicaid expansion, State Plan Amendment (SPA) optional family planning eligibility groups, and Medicaid section 1115 waiver demonstration projects have increased eligibility for family planning services. The primary outcome was the state level annual reported rate of CS per 100,000 live births. We assessed two binary policy “treatments” of interest: 1) whether a state had adopted Medicaid expansion and 2) whether a state had adopted either Medicaid expansion, a family planning waiver, or a family planning SPA. We estimated dynamic and overall treatment effects using a novel difference-in-differences estimator that accounted for variation in treatment timing. Sensitivity analyses controlled for state-level characteristics including education, poverty, unemployment, race, ethnicity, migration, and population size change.

**Data Sources and Analytic Sample:** Data were collected from CDC surveillance reports and 1-year American Community Survey files from 2008-2022.

**Principal Findings:** Pre-treatment trends in CS outcomes were similar between states that implemented the Medicaid policies and those that did not. Medicaid expansion alone was associated with a significant reduction of 6.37 (95% CI: 1.94, 10.79) cases per 100,000 live births over the first three years of the policy. Adopting either Medicaid expansion, a family planning SPA, or a family planning waiver was related to a non-significant reduction of 5.06 (95% CI: −3.21, 13.33) cases per 100,000 live births. Significant reductions in CS were also found within the first year of policy adoption.

**Conclusions:** This analysis found Medicaid expansion policies were associated with reductions in CS rates. Increasing access to family planning services through Medicaid programs might reduce CS even shortly after policy adoption.

## Introduction

Addressing the rise in rates of congenital syphilis (CS) is an urgent public health priority.^1^ CS occurs when syphilis is vertically transmitted to a fetus during pregnancy and is prevented through diagnosis and treatment of syphilis before or early during pregnancy. Untreated syphilis can have severe health consequences, including neurologic changes and permanent vision and hearing loss for infants in addition to impacting pregnancy outcomes.^2^ Reported cases of CS in the United States have increased an alarming 10-fold over the past decade to 3,755 cases in 2022.^1,3^ CS is associated with spontaneous abortion, stillbirth, infant death, pre-term delivery, low birth weight, and long-term health impacts for surviving infants.^1,4^ Further, hospitalizations attributable to CS have high direct medical costs.^5,6^

A variety of factors may be contributing to the syphilis and CS epidemics, and studying their social, behavioral, and clinical determinants might yield critical information for controlling the rise of syphilis and CS.^1,2^ First, rates of CS are closely related to rates of primary and secondary (P&S) syphilis in women of reproductive age (15-44 years), so factors contributing to the rise in P&S syphilis in this population are relevant.^3^ There is evidence of increases in sexual behaviors that are associated with STIs: condom use has fallen among some groups in recent years,^7,8^ and there may be changes in safe-sex behaviors in response to falling HIV mortality. ^9^ Changes in technology may be altering the composition of sexual networks,^10^ which are a critical component of sexually transmitted infection (STI) transmission. Trends in opioid use, methamphetamine use, and transactional sex may also be contributing to the rise in syphilis in the U.S.^11,12^ Further, healthcare disruptions during the COVID-19 pandemic likely led to reductions in STI services.^13^

Since 2010, substantial changes to the U.S. healthcare landscape, including family planning services (e.g., contraception, STI services, and preconception services), also have possible implications for syphilis and CS. Starting in 2014, as part of the Affordable Care Act (ACA), many states and the District of Columbia (D.C.) expanded eligibility for full-benefit Medicaid, i.e., comprehensive insurance that covered all mandatory Medicaid benefits.^14^ Medicaid, the primary funding source for family planning services for people with low incomes, has established minimum federal standards for family planning services and supplies, with states able to set eligibility levels and determine the amount, duration, and scope of covered benefits within broad federal parameters.^15,16^ The ACA required many group and individual health plans (i.e., private insurance) and Medicaid plans to cover preventive services without cost sharing, including FDA approved contraceptives, STI and HIV screenings, well-woman preventive visits, breast and cervical cancer screenings, and HPV vaccination.^17,18^

In addition to full-benefit Medicaid expansion, limited-benefit Medicaid policies have extended eligibility for family planning specific care since the 1990s. Medicaid section 1115 family planning demonstration waivers are time-limited programs that extend coverage only for family planning services to additional people low incomes who do not qualify for full-benefit Medicaid. Further, the ACA included a provision for state plan amendments (SPAs) that enabled states to establish family planning expansion programs by permanently amending their Medicaid state plan without the need for federal renewal.^19^ Combined, changes in these Medicaid policies led to substantial shifts in eligibility for family planning services through Medicaid during 2008-2022.^20^ Subsidies for directly purchasing private health insurance have also made healthcare more affordable for women with low incomes during this period.^14^ Public STI and family planning clinics form additional key infrastructure for the family planning safety net in the U.S., but recent declines in other sources of federal and state funding to these clinics may have made revenue from Medicaid relatively more important.^21,22^

Despite gains in health insurance coverage following the implementation of the ACA,^23^ gaps in access persist.^24^ Increases in eligibility for Medicaid or subsidized health insurance do not always translate into enrollment and utilization.^20^ Persistent disparities and gaps in healthcare access likely mean continued missed opportunities to prevent CS especially among certain groups. Racial, ethnic, and income disparities in reported rates of syphilis have been documented, and rates are often increasing faster in areas with higher levels of poverty.^25,26^

The objective of this analysis was to assess the relationship between the adoption of Medicaid expansion policies and rates of CS in the United States. Numerous studies have explored the impact of Medicaid expansion on a variety of health and economic outcomes.^27^ However, the existence of limited-benefit family planning waivers and SPAs might complicate the relevant timing of policy changes for sexual and reproductive health outcomes affected by access to family planning services. For example, some states that expanded full-benefit Medicaid in 2014 already had waivers or SPAs in place that extended access to family planning services, meaning there was not have been a significant increase in eligibility for these services when the state expanded Medicaid. Building on previous work that characterized the complex policy landscape,^20^ we use quasi-experimental designs to analyze Medicaid’s impact on CS. Our work highlights how adoption of Medicaid expansion policies might prevent CS and further costly downstream health outcomes.

## Methods

To assess how Medicaid expansion policies impact reported rates of CS, we leveraged policy variation among states between 2008 and 2022 and implemented a novel estimator of difference-in-difference (DiD) that accounts for variation in treatment timing. There were two primary policy changes, or “treatments,” of interest. Under the first, Medicaid expansion alone (ME), states were considered treated if they had implemented Medicaid expansion. Under the second, Medicaid expansion plus limited family planning policies (ME+LFP), states were considered treated if any of Medicaid expansion, a family planning waiver, or a family planning SPA were in place at any point in a given year. The treatment variable is binary where one means a state is treated in that year, and zero otherwise. Note that our treatment variables measure policy adoption, not actual healthcare utilization or access. States were considered treated based on policy implementation, regardless of subsequent enrollment levels or service utilization. This shifts the timing of treatment for states that have a limited-benefit family planning program in place at some point during the study period. In the analytic dataset, these treatment variables were coded as binary indicators taking the value of one if a state was treated during a given year and zero otherwise. States that experienced policy changes (e.g., ending a waiver program or transitioning between policy types) were coded according to their treatment status in each specific year. For the ME+LFP analysis, 26 states experienced treatment changes during the study period, with most changes involving transitions to Medicaid expansion (n=16), followed by SPAs (n=7) and waivers (n=3). One state experienced multiple treatment changes during the study period.

DiD estimators use two types of comparisons to calculate treatment effects. First, outcomes within a state before treatment onset are compared to outcomes once treatment has begun. The change within a treated state is then compared to the concurrent change in states that were not treated during that same period (i.e., the controls). The controls included states during any years they were untreated. Specifically, states served as controls if they either: 1) never implemented the policy during the study period, or 2) had not yet implemented the policy in a given year.

There were two effect lengths highlighted in this analysis: the effect one year after the start of treatment and the overall effect, which used three years of post-treatment data unless otherwise noted. Three years was selected for the overall effect after treatment initiation for several reasons. First, expanding the treatment length would result in fewer states meeting the inclusion criteria, reducing the sample size as well as the precision of the estimates. For example, using a four-year window would exclude an additional three states that expanded Medicaid after 2019. Second, a three-year window provides sufficient time to observe policy effects while minimizing the influence of other concurrent policy changes or external factors that could affect outcomes. We therefore dropped treated states with fewer than three years of treated status in our main specification (n=2) to avoid changes in the effect due to compositional changes from differences in the set of states used to determine the 1-year versus overall effects. States that were always treated (e.g., had a waiver in place starting in 2008, n=21) were systematically dropped from the ME+LFP analysis because they do not have pre-treatment data and are not considered valid comparators. The preferred estimator for the DiD models was proposed by de Chaisemartin and D’Haultfœuille (dCDH) and is well suited for this analysis because it accommodates treatment switching and semi-continuous treatments, which were used in some sensitivity analyses.^28^

The primary model outcome is reported cases of CS per 100,000 live births. We hypothesized two basic mechanisms through which family planning services might prevent cases of CS. First, STI testing and treatment might reduce syphilis among women of reproductive age who are pregnant or who may later become pregnant. Second, affordable access to contraception might reduce unintended pregnancies, which may in turn reduce cases of CS. To test these hypotheses, we estimated models using two additional outcomes: 1) the rate of primary and secondary (P&S) syphilis among women aged 15-24 years, and 2) total births per 1,000 women aged 15-19 years. Syphilis data were retrieved from the National Center for HIV, Viral Hepatitis, STD, and Tuberculosis Prevention (NCHHSTP) AtlasPlus and live birth outcomes from CDC Wonder.^29,30^ Nearly half of reported STIs occur among adolescents and young adults aged 15-24 years,^3^ and this group is more likely to be enrolled in Medicaid.^31^ Live birth data were restricted to women aged 15-19 years because previous research found large effects of Medicaid family planning policies within this group.^32^

We conducted a variety of sensitivity analyses. First, in place of a binary treatment indicator, the income eligibility threshold for women without children (with the dCDH estimator) was used. Next, the proportion of adult reproductive aged women (aged 18-44 years) who were eligible for family planning services through Medicaid was estimated. The calculation of this variable is detailed in Williams et al. (2024).^20^ The effect of the size of the eligible population was estimated using an instrumental variables estimator with year and state fixed effects, following Currie and Gruber (1996).^33^ The instrumental variable for a state’s estimated eligible population was the national proportion of women that would be eligible for Medicaid family planning if the state-level eligibility criteria were applied nationally. This strategy accounts for the possibility that state decisions to implement policies were in part based on state-specific demographic or economic trends, which would otherwise bias the estimated relationship. A one-year lagged version of this estimator was also utilized. We did not estimate a model with a lagged effect using the main specification because the dCDH estimator compares all other outcomes to the period just before initiation of treatment. If there is any effect in the first year, then that becomes the baseline in a lagged model and reduces estimated effects in later years. Next, additional models were estimated that included covariates, included states that had been recently (i.e., less than three years) treated, used four years of post-treatment data, or implemented the Callaway and Sant’Anna estimator.^34^ For sensitivity analyses of the mechanism models, we estimated models with the following additional outcomes: P&S syphilis in women of reproductive age (15-44 years), overall live births (total births per 1,000 reproductive aged women), and the proportion of births to people with incomes below the poverty level. Analyses were conducted using Stata 18 and R version 4.4.0.

The instrumental variable models included a variety of control variables: the proportion of adult reproductive age women completing at least one year of higher education, mean income of adult reproductive age women as a proportion of the federal poverty level, the proportion of adult reproductive age women in households that received benefits from the Supplemental Nutrition Assistance Program (SNAP), the proportion of the state population that is unemployed, the proportion of the adult reproductive age female population that identifies as non-Hispanic Black, the proportion of the adult reproductive age female population that identifies as non-Hispanic White, the proportion of the adult reproductive age female population that identifies as Hispanic, the proportion of the state population that migrated from out of state in the past year, the annual percent change in state population, the proportion of the state population that is female, the proportion of the female population that is between the ages of 15 and 24 years, and the proportion of the female population that is between the ages of 25 and 44 years. Covariate data were derived from American Community Survey data accessed via IPUMS.^35^ Some covariate data were restricted to adult reproductive age women (i.e., 18-44 years) because it mirrors eligibility for some government programs, including family planning waivers in some states. The main dCDH specifications did not include covariates because 1) the assumptions of parallel trends appeared to be satisfied without covariates (i.e., we observed parallel trends in CS rates among treatment and non-treatment states prior to the onset of treatment), and 2) not all covariates could be used in some treatment years because of the relatively small number of control observations.^28^

## Results

Twenty-seven states (including D.C.) expanded full-benefit Medicaid in 2014 (Supplemental Figure 1). Three more states expanded in 2015, two in 2016, two in 2019, and three in 2020. Oklahoma and Missouri expanded in 2021 but were not included in the main estimates because they did not have three years of post-treatment data at the time this study was conducted. Thus, there were 37 states that switched their treatment status under the ME treatment definition. For the ME+LFP treatment, 26 states switched their treatment status during the study period (Supplemental Figure 2). The relevant treatment change was Medicaid expansion in 16 states, SPAs in 7 states, and waivers in 3 states. Missouri and D.C. had state funded programs that provided similar levels of family planning benefits prior to Medicaid expansion and were thus designated as always-treated.^20^ There were fewer total treatment changes under the ME+LFP definition because many expansion states already had waivers in place prior to 2008 and therefore did not experience a change in treatment status. For example, Illinois had a family planning waiver in place prior to 2008. When they expanded Medicaid in 2014, there was no change in the ME+LFP treatment variable because they already had a Medicaid policy in place for family planning services.

The mean rate of CS across states during 2008-2022 was 21.98 cases per 100,000 live births. There were on average 6.38 cases of P&S syphilis per 100,000 women aged 15-44 years. Summary statistics for additional outcome and control variables are presented in Table 1. There were no significant differences in CS rates between treated and untreated states prior to Medicaid expansion (Figure 1). Using the main model specifications, Medicaid expansion alone (ME) was associated with a reduction of 4.59 (95% CI: 1.62, 7.56) cases per 100,000 live births after one year and 6.37 (95% CI: 1.94, 10.79) cases per 100,000 live births overall (Table 2). Adopting Medicaid expansion, a family planning waiver, or a SPA (ME+LFP) was associated with reductions of 3.37 (95% CI: 0.23, 6.51) cases per 100,000 and 5.06 (95% CI: −3.21, 13.33) cases per 100,000 after one year and overall, respectively.

**Figure 1:**
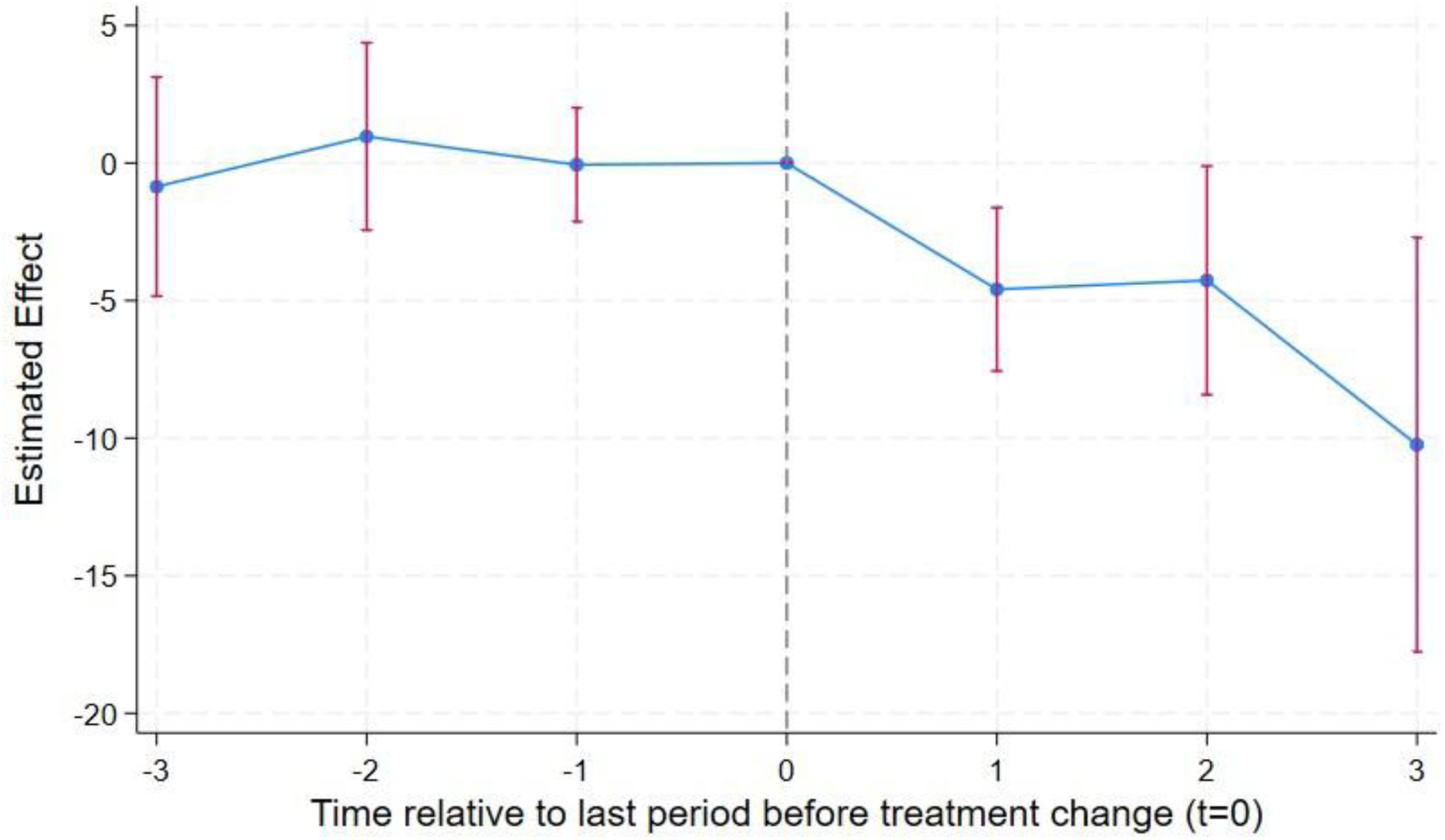
Event Study - Relationship between Medicaid Expansion and the Rate of Congenital Syphilis

**Table 1:** Mean and Standard Deviation of Outcome Variables and Covariates.

| Variable | Mean | Standard Deviation |
| --- | --- | --- |
| <u>Outcomes</u> |  |  |
| CS rate | 21.98 | 41.68 |
| Primary and secondary syphilis rate (ages 15-44) | 6.38 | 11.81 |
| Primary and secondary syphilis rate (ages 15-24) | 6.35 | 10.55 |
| Total births per 1,000 women (ages 15-44) | 62.01 | 7.35 |
| Total births per 1,000 women (ages 15-19) | 24.39 | 11.57 |
| Proportion of births to women below the FPL | 7.56% | 1.98% |
| <u>Alternative Treatment Variables</u> |  |  |
| Income eligibility threshold as percentage of FPL (for women without children) | 138.92 | 95.84 |
| Eligible Proportion | 37.21% | 18.02% |
| <u>Covariates</u> |  |  |
| Proportion receiving SNAP benefits* | 16.85% | 5.1% |
| Proportion completing at least 1 year of higher education* | 62.67% | 5.4% |
| Income as a proportion of the FPL* | 2.90 | 29.65 |
| Proportion migrating from out of state in past year | 3.54% | 1.34% |
| State unemployment rate | 4.97% | 1.78% |
| Proportion of population that is female | 50.60% | 0.85% |
| Population growth | 0.67% | 0.85% |
| Proportion identifying as Black, Non-Hispanic* | 12.84% | 11.17% |
| Proportion identifying as White, Non-Hispanic* | 67.64% | 16.16% |
| Proportion identifying as Hispanic* | 12.91% | 11.14% |
| Proportion of the female population aged 15-44 years | 13.13% | 0.98% |
| Proportion of the female population aged 25-44 years | 25.81% | 2.06% |

\*Among women aged 19-44 years
CS: congenital syphilis
SNAP: supplemental nutrition assistance program
FPL: Federal Poverty Level

**Table 2:**
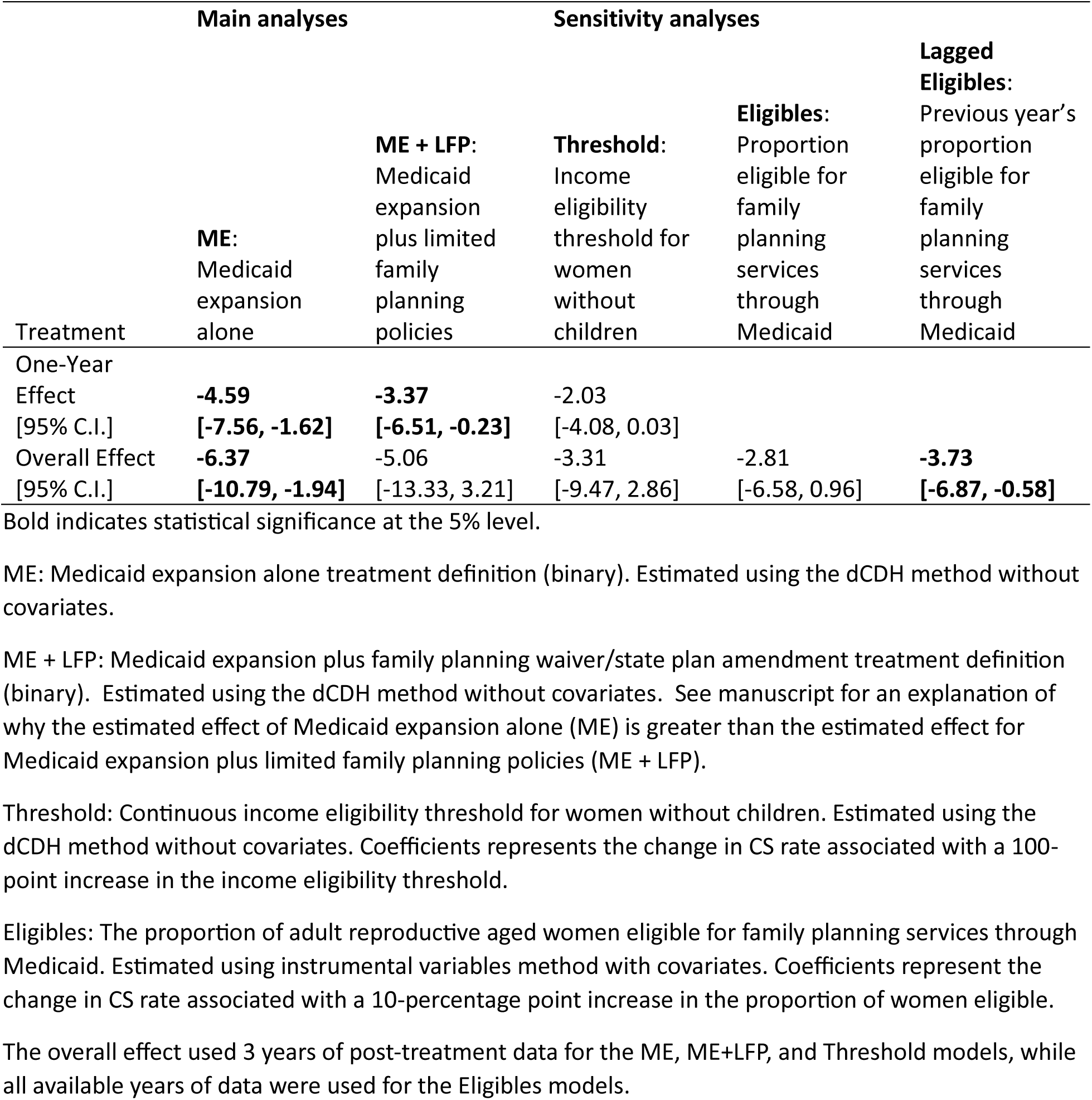
Estimated change in the number of congenital syphilis cases per 100,000 live births after implementation of Medicaid policies.

Other treatment definitions resulted in similar CS rate reduction estimates, but only the lagged eligible proportion estimate was statistically significant (3.73 cases per 100,000 overall effect, 95% CI: 0.58, 6.87). For secondary outcomes, there was some evidence of a reduction in P&S syphilis in women ages 15-24 (2.36 cases per 100,000 overall effect, 95% CI: 0.74, 3.97) and no evidence of an effect on live births among women ages 15-19 (Table 3). The main results were generally robust when using alternative models and specifications (Supplemental Table 1). The one-year effects were statistically significant in all but one the CS outcome models, as were three of the five overall effects. The two alternative live birth outcome models yielded no significant effect estimates.

**Table 3:** Estimated change in P&S syphilis and live births after implementation of Medicaid policies.

|  | <u>P&S Syphilis Rate (15-24)</u> |  | <u>Live Birth Rate (15-19)</u> |  |
| --- | --- | --- | --- | --- |
|  | ME:<br>Medicaid<br>expansion alone | ME + LFP:<br>Medicaid expansion plus<br>limited family planning<br>policies | ME:<br>Medicaid<br>expansion alone | ME + LFP:<br>Medicaid expansion plus<br>limited family planning<br>policies |
| One-Year Effect | **-1.66** | -0.76 | 0.07 | -0.07 |
| [95% C.I.] | **[-3.11, -0.21]** | [-2.37 0.84] | [-0.46, 0.60] | [-0.64, 0.49] |
| Overall Effect | **-2.36** | -1.54 | 0.14 | -0.26 |
| [95% C.I.] | **[-3.97, -0.74]** | [-4.93, 1.86] | [-0.62, 0.90] | [-1.16, 0.63] |

Syphilis and live birth rates are among women. Bold indicates statistical significance at the 5% level. The overall effect included 3 years of post-treatment data.
P&S: primary and secondary stages
ME: Medicaid expansion alone treatment definition. Estimated using the de Chaisemartin and D'Haultfœuille (dCDH) method without covariates.
ME + LFP: Medicaid expansion plus family planning waiver/SPA treatment definition. Estimated using the dCDH method without covariates.

## Discussion

This analysis found robust evidence that Medicaid policies that increased eligibility for family planning services were associated with declines in the rate of CS. Medicaid expansion alone was associated with a significant reduction of over 6 cases per 100,000 live births compared to rates in non-expansion states, equivalent to 29% of the mean rate of CS across states from 2008-2002. Medicaid expansion plus the extension of family planning services through limited-benefit waivers or SPAs was associated with a reduction of over 5 cases per 100,000 live births over three years of implementation, but the estimate was not significantly different from zero. The difference in precision between the ME and ME+LFP results does not necessarily mean that the family planning specific polices were themselves ineffective, rather that the effect signal from the Medicaid expansion analysis, which didn’t account for policies in place prior to expansion, may have been stronger. There are a variety of reasons that may lead to less precise effect estimates in our data. Limited-benefit waivers or SPAs, including those extending eligibility for people who are pregnant or family planning, might be less publicized and well-known. Medicaid expansion occurred in many states simultaneously and benefited from large scale outreach campagins.^36^ The heightened awareness might have led to more enrollment and service utilization. This is consistent with previous work showing that even though higher pregnancy eligibility limits were in place prior to implementation, Medicaid expansion improved timeliness and adequacy of prenatal care.^37^ In addition, in some states family planning program outreach was focused on existing clinic users.^19^ This focus might have resulted in a change in payer rather than increased utilization for those patients, although the influx of Medicaid dollars might have expanded clinic capacity for future clients.^19^ In addition, full-benefit Medicaid expansion included access to additional preventive services beyond family planning, which could have additional contributions to preventing CS.

The estimated effect on CS was statistically significant in the first year the policies were implemented and grew slightly in magnitude over the first three years after policy adoption. Outreach and health communication strategies in expansion states might have increased awareness of the upcoming policy change and allowed people to quickly utilize the new benefits.^36^ Given that diagnosing and treating syphilis prior to pregnancy and early during pregnancy can be quickly accomplished when people present to care,^1,2^ it is reasonable that increasing access to clinical services that include syphilis management would be able to prevent CS after just one year. The growing effect size over time is unsurprising, as the prevention activities could impact pregnancy outcomes months to years after the service was received. The instrumental variable estimates, using eligible population sizes, yielded additional evidence to support a growing effect over time. In those sensitivity analyses, the estimated effect was larger and more precise when the treatment variable was lagged one year.

We found that Medicaid expansion was associated with lowered P&S syphilis in women of reproductive age, but we found no evidence of an association with live births. Lowering background rates of P&S syphilis in women reduces the incidence of CS and its sequelae. Additionally, while access to affordable contraception and preconception care could theoretically affect CS incidence in multiple ways, including by reducing unintended pregnancy, our analysis found evidence only for reductions in P&S syphilis rates as a potential mechanism. Previous evidence on the effect of limited family planning or full-benefit Medicaid expansion on live birth outcomes has been mixed,^27^ but some studies did find reductions in births following the policy implementation.^32,38,39^

This analysis omits additional policy complexity that might be relevant for syphilis and CS outcomes. States have phased out using waivers in favor of SPAs, but this analysis did not distinguish between those two limited-benefit mechanisms and therefore might miss any potential benefits from switching to a SPA. Medicaid eligibility extensions for people who are pregnant have been in place for decades. However, pregnancy eligibility was fairly constant and did not vary substantially across states during the time period of this analysis, so it should not impact the relevant timing of treatment in this analysis. In addition, the ACA instituted marketplaces which may have facilitated access to insurance, required private insurance plans cover certain family planning services, and extended dependent insurance coverage until age 26. However, these changes were applied to all states simultaneously, so they should not affect the DiD estimates. The Title X Family Planning Program, a domestic federal program dedicated to ensuring access to high-quality, client-centered family planning care for millions of people with low incomes or who are uninsured, may interact with Medicaid funding in complex ways but was omitted from the current analysis.^22^ Our treatment variables also capture policy adoption rather than actual healthcare access or utilization. While policy implementation is necessary for expanding access to care, other barriers such as awareness, enrollment processes, provider availability, and other factors can affect whether eligible individuals ultimately receive services. Our estimates thus represent the intent-to-treat effect of these policies rather than the effect of actually receiving services. While our 3-year post-treatment window captured significant effects, longer-term impacts may differ. Additionally, the complex nature of state policy changes, including transitions between different types of family planning coverage and occasional policy reversals, required simplifying assumptions in our treatment coding that may not fully capture all policy nuances. Finally, there is variation in state laws regarding the timing and frequency of required screening for syphilis during pregnancy.^40^

This analysis provides new evidence on the potential impact of adopting Medicaid expansion policies on CS prevention. Policymakers and public health professionals might consider these findings when considering intervention alternatives for addressing the syphilis and CS epidemics. States could consider the potential benefit of leveraging their Medicaid programs to reduce CS rates through expansion of health insurance coverage, especially to people with low incomes who might not be able to otherwise afford care, as well as the availability of low-cost family planning services. There are promising new mechanisms for increasing eligibility for services through Medicaid that might be beneficial in CS prevention among people who have limited access to social and medical services, including SPAs that expand 12-month coverage during the postpartum period and section 1115 waivers that address health-related social needs.^41^ Further research might be beneficial to understand the complex relationships between legislated policies and subsequent enrollment and utilization of services relevant for addressing syphilis and CS. Mediating factors, such as health care workforce capacity, education efforts, and health, also play important roles in the implementation and effectiveness of syphilis and CS prevention policies.

## Data Availability

All data sources were publicly available when this study was conducted. Health outcome data came from the US Centers for Disease Control and Prevention (https://gis.cdc.gov/grasp/nchhstpatlas/main.html; https://wonder.cdc.gov/). Additional economic and demographic data were downloaded from IPUMS USA (https://usa.ipums.org/usa/).

---

**Supplemental Table 1:**
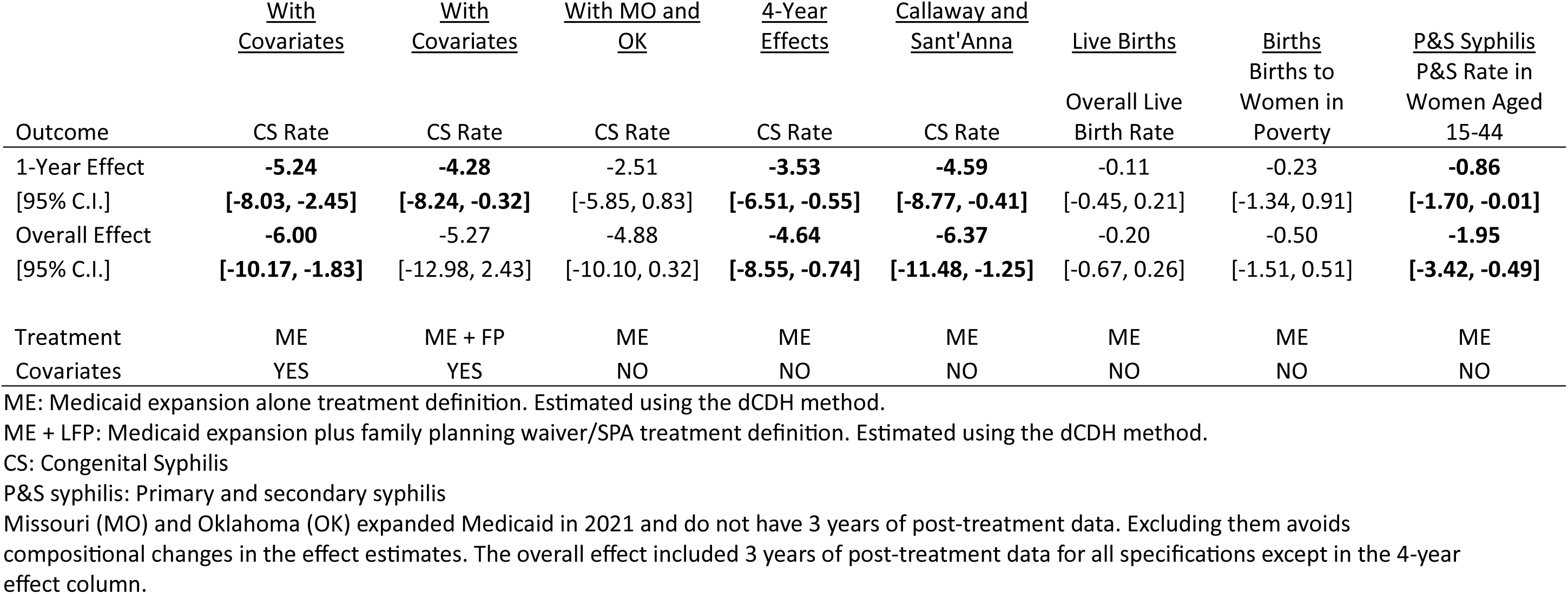
Alternative Models and Specifications for the Relationship between Medicaid Policies and CS, P&S Syphilis, or Live Births.

**Supplemental Figure 1:**
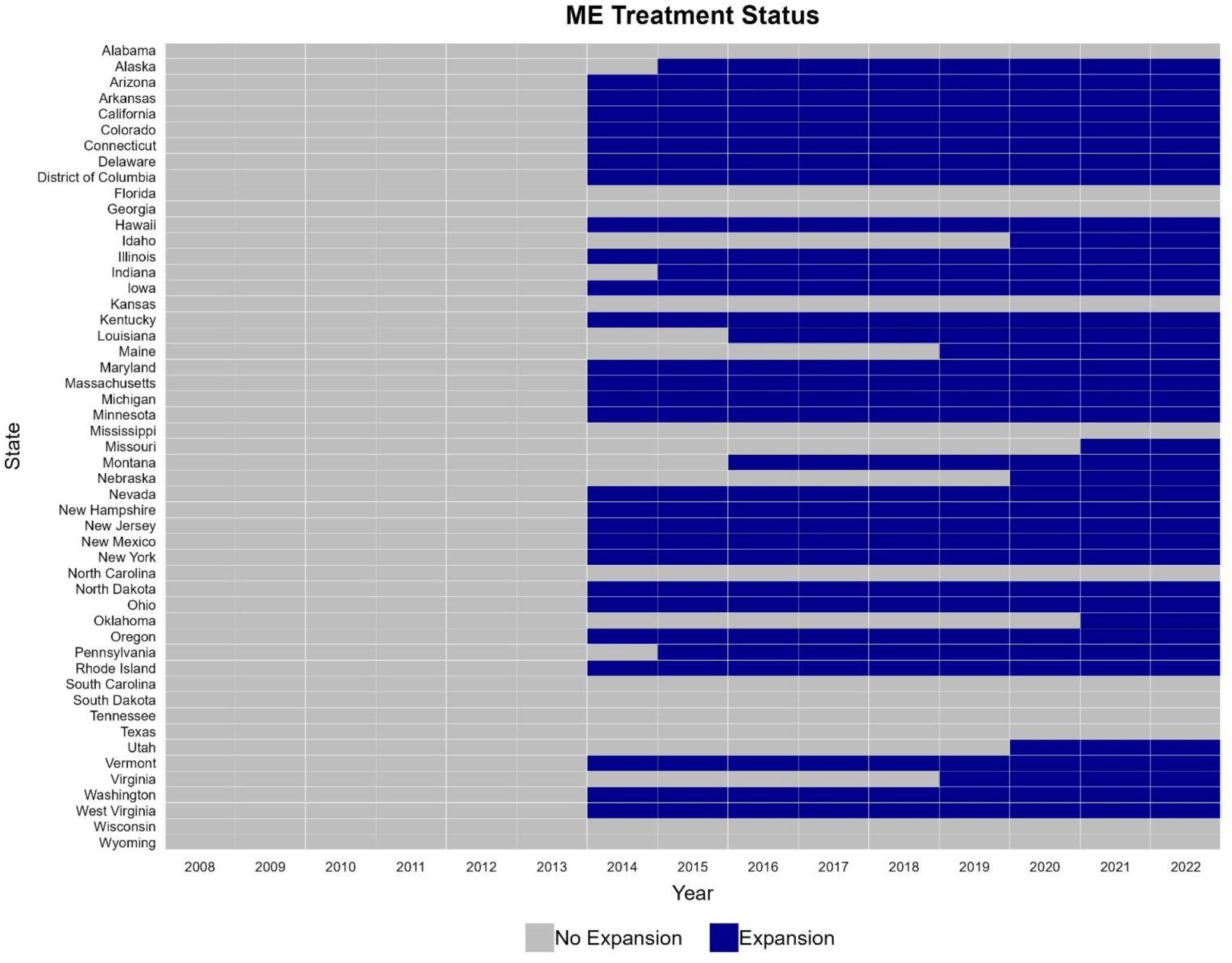
Timing of Medicaid Expansion by State ME: Medicaid expansion alone treatment definition.

**Supplemental Figure 2:**
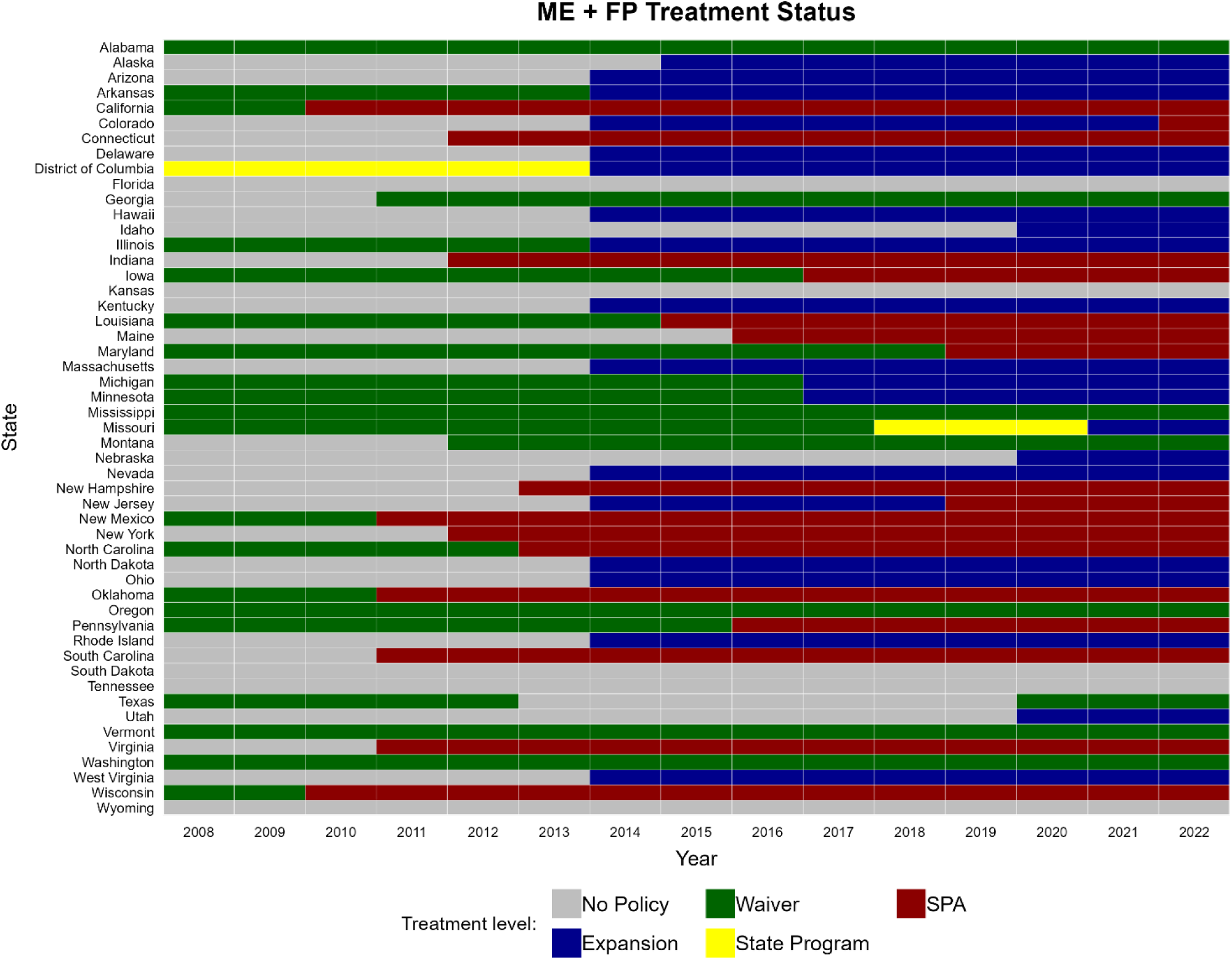
Timing of Medicaid Family Planning Waivers, Medicaid Family Planning State Plan Amendments, and Full-Benefit Medicaid Expansion, by State ME + LFP: Medicaid expansion plus limited-benefit family planning waiver/state plan amendment (SPA) treatment definition. When a state has overlapping policies, the color refers to the first policy implemented.

**Supplemental Figure 3:**
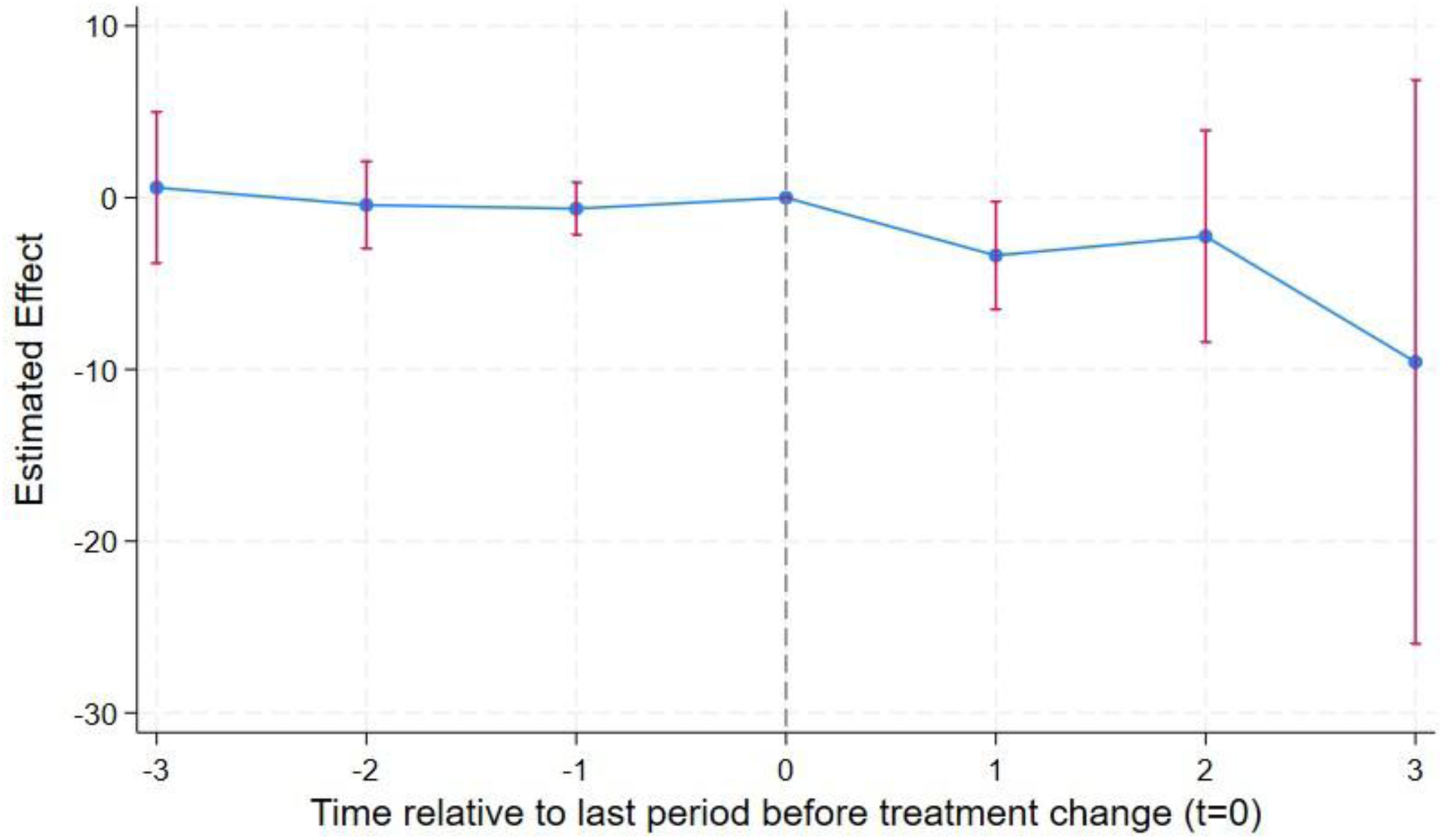
Event Study - Relationship between Medicaid Expansion and Limited-Benefit Family Planning Waivers/SPAs and the Rate of Congenital Syphilis

**Supplemental Figure 4:**
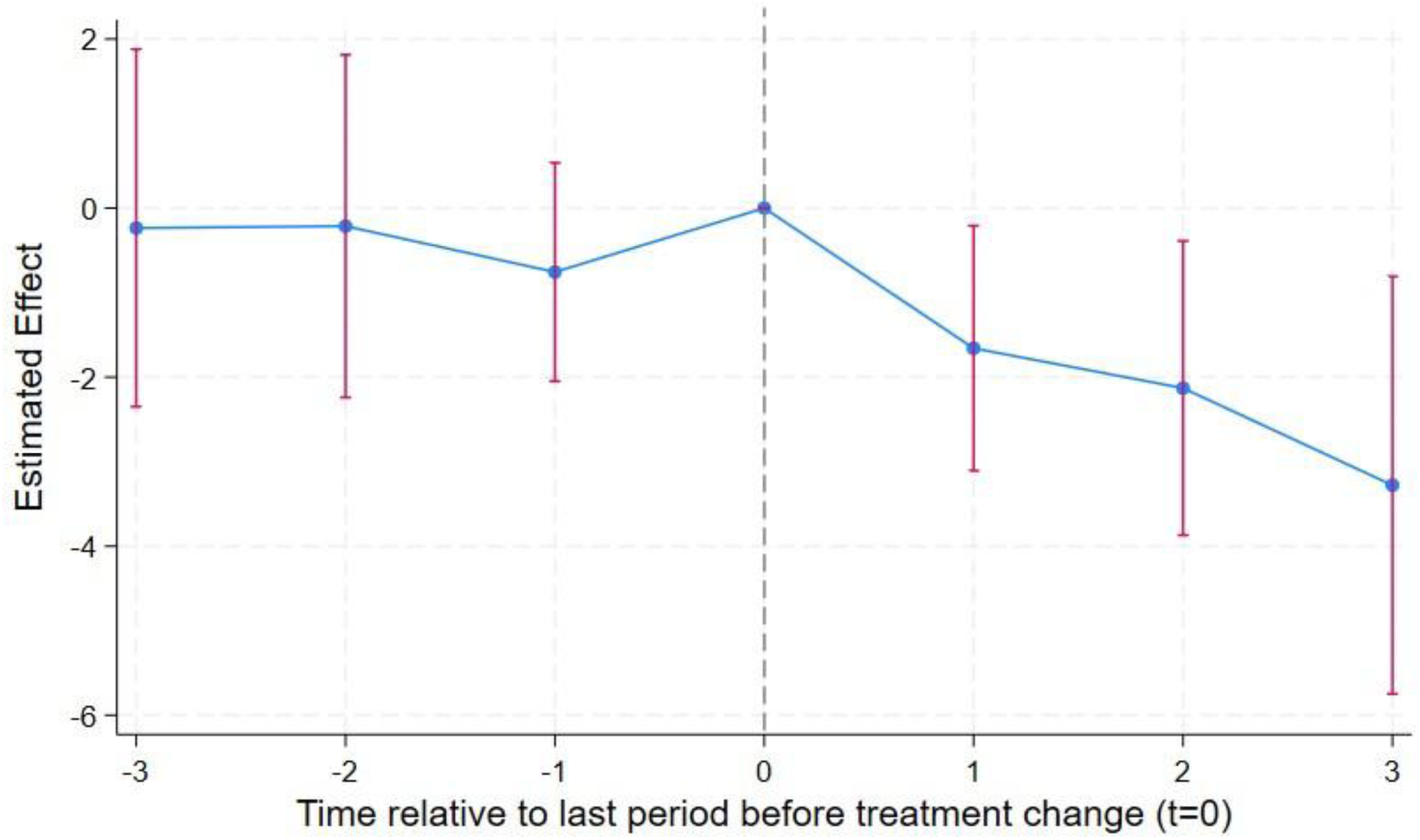
Relationship between Medicaid Expansion and the Rate of Primary and Secondary Syphilis among Women Aged 15-24

## Notes

### Competing Interest Statement

The authors have declared no competing interest.

### Funding Statement

No external funding was received for this study.

### Author Declarations

All data sources were publicly available when this study was conducted. Health outcome data came from the US Centers for Disease Control and Prevention (https://gis.cdc.gov/grasp/nchhstpatlas/main.html; https://wonder.cdc.gov/). Additional economic and demographic data were downloaded from IPUMS USA (https://usa.ipums.org/usa/). Policy details were described in a previous study (https://pubmed.ncbi.nlm.nih.gov/39439245/)

### Summary of Updates

Author contact information was updated.

